# An Automation Framework for Clinical Codelist Development Validated with UK Data from Patients with Multiple Long-term Conditions

**DOI:** 10.1101/2024.09.25.24314215

**Authors:** A. Aslam, L. Walker, M. Abaho, H. Cant, M. O’Connell, A. S. Abuzour, L. Hama, P. Schofield, F.S. Mair, R.A. Ruddle, O. Popoola, M. Sperrin, J.Y. Tsang, E. Shantsila, M. Gabbay, A. Clegg, A.A. Woodall, I. Buchan, S. D. Relton

## Abstract

**Background:** Codelists play a crucial role in ensuring accurate and standardized communication within healthcare. However, preparation of high-quality codelists is a rigorous and time-consuming process. The literature focuses on transparency of clinical codelists and overlooks the utility of automation.

**Method and Automated Framework Design:** Here we present a Codelist Generation Framework that can automate generation of codelists with minimal input from clinical experts. We demonstrate the process using a specific project, DynAIRx, producing appropriate codelists and a framework allowing 1future projects to take advantage of automated codelist generation. Both the framework and codelist are publicly available.

**Use-case: DynAIRx:** DynAIRx is an NIHR-funded project aiming to develop AIs to help optimise prescribing of medicines in patients with multiple long-term conditions. DynAIRx requires complex codelists to describe the trajectory of each patient, and the interaction between their conditions. We promptly generated **≈**200 codelists for DynAIRx using the proposed framework and validated them with a panel of experts, significantly reducing the amount of time required by making effective use of automation.

**Findings and Conclusion:** The framework reduced the clinician time required to validate codes, automatically shrunk codelists using trusted sources and added new codes for review against existing codelists. In the DynAIRx case study, a codelist of **≈**9600 codes required only 7-9 hours of clinician’s time in the end (while existing methods takes months), and application of the automation framework reduced the workload by **>**80%.

## 1 Introduction

In recent years, there has been an increasing reliance on Electronic Health Records (EHRs) to study the health and care of large patient populations. Health systems around the world rely increasingly on the analysis of EHR data to plan and manage the quality of their services - a Population Health Management^1^ (PHM) requirement. To perform these analyses, a critical but often overlooked step is the creation of “codelists” to process the raw patient record into a form suitable for analysis.

The patient record is a collection of coded events (typically using clinical terminologies such as ICD or SNOMED mapped to capture diagnosis, medications, procedures, referrals etc) [1]. Each SNOMED code represents a specific diagnosis, symptom, or treatment and can have multiple variants. For example, SNOMED code “195967001” is Asthma and “281239006” is Exacerbation of Asthma. A codelist groups a set of codes into a clinical concept at the correct level of detail to answer a given research question; in the example above these might fall under a general asthma codelist, or we may need to separate out primary asthma diagnosis from worsening symptoms (depending upon the research question).

These codelists play a crucial role in ensuring accurate and standardized communication within healthcare provision, commissioning, and research. Efforts to be transparent and share codelists such as OpenCodelists ^2^ are welcome additions, but there will always be a need to create new codelists. As demonstrated above, codelists are intimately linked to the research or commissioning question, and the underlying set of SNOMED codes is regularly updated with new additions, meaning that codelists cannot remain static in perpetuity.

Construction of high-quality codelists involves a range clinical, technical, and informatics expertise, meaning it can become a time-consuming process. In spite of the importance of codelists, they are often constructed or updated haphazardly, without any clear guidance or protocol. In this work we proposed a Codelist Generation Framework which derives a process for building codelists using automation where possible to reduce the amount clinical effort required whilst retaining high-quality. We use the ongoing DynAIRx project^3^, focused on multimorbidity, as a case study to show the impact of the framework, and release the code required to implement our framework as open source software.

The resulting framework makes use of trusted sources (such as the Quality Outcomes Framework [2] and CALIBER [3, 4]) and automation to reduce the requirement for clinical expertise. In our case study, a codelist with *≈* 9600 items was compiled using only 7–9 hours of clinicians’ time by employing the proposed framework, and more than 80% of the codes were generated and validated using the framework before clinical validation.

This paper aims to provide a transparent generalized codelist development framework - demonstrated via application to the DynAIRx project - to semi-automate this time-consuming process. Software to enable use of this framework and the resulting DynAIRx codelists are released for public use. The main contributions of this work can be summarized as follows:

- Design of a Codelist Generation Framework, applicable to any codelist generation task, that aims to reduce clinical validation effort significantly.
- Generation of large codelists for the DynAIRx case-study, applicable to cohorts of multiple longterm conditions (multimorbidity) on multiple medicines (polypharmacy)
- Comprehensive evaluation of a codelist generated using the proposed framework including a reduction in clinicians’ workload in generating and validating codes.
- Releasing codelists and making the Generalised Codelist Automation Framework “GCAF” (Python Repository) publicly available for codelist generation.

The remainder of the paper is organized as follows. A brief overview of existing techniques with background are presented in Section–2. The proposed “Generalised Codelist Automation Framework (GCAF)” with details of design, implementation, and examples for different phases are discussed in Section–3. A case-study utilising the framework to generate codelists for the DynAIRx project is presented in Section–4. A comparison of the resulting DynAIRx codelist to common alternatives, and our learning from the process are captured in Section–5. Finally, concluding remarks and avenues for future work appear in Section–6.

## 2 Background

This section aims to provide a comprehensive overview of work in codelist generation to-date. It is divided into three subsections. First, we summarise the existing systematic reviews on codelist development, focusing on the strengths, and challenges they highlight. Second, we describe the recommended best practice with regards to codelist development, and note the issues around automation and reproducibility that motivated this work. Finally, we introduce existing codelists that are commonly used in multimorbidity research.

Codelists can consist of different types of codes like SNOMED, ICD, Read and other ontologies. When using electronic healthcare records (EHRs), clinicians store data about a patient using a standard ontology, commonly SNOMED, CTV2, or CTV3 in a primary care setting, ICD or SNOMED in a secondary care setting, and DMD codes for medications. Each code represents a specific diagnosis, symptom, test, or treatment and can have multiple variants. For example, SNOMED code “195967001” is Asthma and “281239006” is Exacerbation of Asthma. When using EHRs within a research or commissioning context we often want to group together similar concepts into *codelists* that indicates someone has asthma, or any other condition of interest. The ontology within which clinicians record this information has evolved over time (and continues to evolve regularly); meaning that codelists also need to be regularly rebuilt to capture this evolving system. The increased interest in research and policies to tackle the multimorbidity and polypharmacy arising from aging populations poses a major challenge for codelist generation due to the size and complexity of these ontologies.

Codelist publications typically refer to academic or research papers that focus on the development, implementation, or analysis of codelists in various fields such as healthcare, bioinformatics, data science etc. It is reported in the literature [5] that crafting high-quality codelists is time-consuming and requires a range of clinical, terminological, and informatics expertise. Various synonyms for codelists are used interchangeably including “value set”, “code set”, “concept set”, and “enumeration”. Another important point they raise is that, despite widespread agreement on the importance of reusability, codelists often suffer from clutter and redundancy, greatly complicating efforts at reuse. When users encounter multiple codes with the same name or ostensibly representing the same clinical condition, it can be difficult to choose amongst them or determine if any differences among them are intentional or due to error.

There is a range of literature on the subject including the definition of codelists [6–11], standardization of methods [12–16] and tools for codelists [17, 18], for assessing codelist quality and terminologies [19–22] [23–25], and for enabling/promoting sharing of codelists for reuse [26–29]. It is clear then that many different codelists are required throughout the healthcare system when using routine datasets and, as they can be problem dependent and time varying, we often need to reuse and adjust existing codelists. At present there is no clear framework for how to do this systematically, or how to leverage automation to decrease the amount of manual effort required during this process. The primary aim of this work is to provide a unifying framework that maximises automation and enables sharing of the construction process.

### 2.1 Codelist Limitations highlighted by Systematic Reviews

Many systematic reviews of codelists found the idea of transparency and reporting of development methods as key requirement. One review of codelists [30] identified codelists related to hypertension that use EHRs and generated recommended codelists. Massen et al. reviewed the literature, providing an extensive summary of codes reported to be used to define hypertension in publications using EHR data. The breadth of codes used to define hypertension varied between studies, leading to selection bias in the resulting research cohorts. They also encouraged a transparent methodology for codelist creation, which is essential for replication and aids in the interpretation of study findings. The framework proposed here has transparency and reproducibility as key elements of the design.

Another review [31] recognized the importance of constructing reliable and reusable codelists. However, the authors found that codelist definitions are rarely transparent and are seldom shared. There is a lack of methodological standards for the management (construction, sharing, revision and reuse) of clinical codelists which needs to be addressed to ensure the reliability and credibility of research. This paper reviewed thirty methodological papers on the management of codelists and provided best practice recommendations for designing and implementation for future studies. The paper emphasised the need for software tools to enable users to easily and quickly create, revise, extend, review, and share codelists.

Subsequent research [32] in the paper “Term sets: A transparent and reproducible representation of clinical code set” also highlights the need for transparency and reproducibility. They also propose the terminology “term sets” (equivalent to codelist) that are findable, accessible, interoperable. This work focused on 31 codelists and released them publicly. However they did not make use of automation to improve the reproducibility and transparency of the development process.

Similarly other research [33–35] also encourages transparency and focuses on creating online repositories which can be used and modified publicly. However, releasing publicly available codes or making them useful widely for EHRs is not sufficient for reproducibility and transparency, it needs to have transparent method of codelist development. Our work addresses these issues by developing an open source toolkit and framework for generating codelists.

### 2.2 Codelists Development Strategies

In this subsection we detail some published procedures for codelist generation, noting that most research does not have a transparent process for reproducibility. The use of UK primary care EHRs for developing codelists has been described in [36]. In that work, codelists were used to estimate the frequency of shortness of breath in a cohort of 28’216 patients within Clinical Practice Research Databank (CPRD) data. Its design is a three-stage process: a priori discussion with clinical experts to look at features of interest, a thorough search for potentially relevant codes using computer software, and clinical agreement via a modified Delphi process (with an “uncertain” category for further sensitivity analysis). Lastly, use same Delphi process to reach consensus between primary care practitioners. There is limited discussion on the time and effort required for codelist generation, the research is focused on reporting of the approach taken. Watson et al. highlighted that codelist generation method is time-consuming, exhaustive, and needs modifications for future EHR studies.

Other strategies for developing clinical codelists have been published, for example [37]. This paper is focused on optimisation of EHR use to describe rheumatoid arthritis in primary care. This paper proposed a methodology to develop “indicator markers” found in patients with early rheumatoid arthritis. They also propose a priori and a posteriori strategies for codelist development. This work discussed an iterative process for constructing codelists. First, a priori indicator markers are produced and, after intermediate steps, the a draft codelist is scrutinized by clinicians. The second, a posteriori, stage of this process involved a further review of the generated codes – though the exact process for reaching clinical agreement was not specified. Although codelists are reusable, they will ultimately need modifications for future projects, and the approach will require extensive involvement of clinical expertise during codelist modification.

### 2.3 Existing Codelists

#### CALIBER

CALIBER ([3, 4]) is the Health Data Research UK (HDR-UK) National Phenotype Library that provides comprehensive codelists for a variety of conditions. CALIBER is an open-access resource led by Spiros Denaxas and provides the research community with information, tools and phenotyping algorithms for EHR data. As the UK National Health Service (NHS) captures huge amounts of clinically coded data, CALIBER is a valuable resource for researchers. However, clinicians sometimes use different codes for the same term in different settings/contexts and therefore using and maintaining the codelist can be challenging. CALIBER also offers algorithms to help infer codes where they are missing, for example a diagnosis code for psychosis if a patient has been prescribed an anti-psychotic medication.

#### electronic Frailty Index (eFI)

Increased interest in looking at the impact of MLTCs on patient outcomes has led to the need for codelists summarising large numbers of conditions. One key example implemented within UK primary care systems and the NICE guidelines is the electronic Frailty Index (eFI). Published in 2016 it contains 1691 SNOMED codes and is used to give a general overview of health in geriatric patients. The eFI2 will be released imminently and contains 7556 SNOMED codes. Both of these are available in multiple ontologies (SNOMED, CTV2, and CTV3) to enable better coverage of the population.

#### OPTIMAL

The OPTIMAL [38] project focuses on improving therapies and AI-assisted clinical management for patients with complex MLTCs. It addresses the challenge of doctors treating diseases individually, often without knowing how treatments for one condition might affect another. By identifying interactions between diseases and treatments, OPTIMAL aims to help clinicians choose therapies that improve outcomes for patients with multiple conditions. This large project Optimal also prepared 30061 codes based codelists with help of clinicians. This motivates the reported research on automated codelist generation by reducing the manual effort required to identify relevant treatments and conditions, improving efficiency and accuracy.

#### AI-MULTIPLY

The AI-MULTIPLY [39] project focuses on understanding the complex interactions between MLTCs and the use of multiple medications (polypharmacy). By analyzing relationships between conditions, treatments, and personal factors, it aims to optimize patient care. These conditions were reviewed by *general practitioners, psychiatrists, geriatricians, gynaecologists, obstetricians, gastroenterologists*, and *dibetologists*. This collaboration between Newcastle University and Queen Mary University highlights the need for accurate condition lists, which are reviewed and refined by healthcare specialists.

In general, codelist development is a key step for research projects to undertake before progressing with the study itself [40–42]. There are numerous existing codelists such as those above and other efforts including the UK Biobank [43] and Cambridge codelists [44]. However, projects often need to create/modify these existing codelists leading to issues of transparency and reproducibility as highlighted in the previous sections.

### 3 The Codelist Generation Framework: Methodology and Automation

In this section we design a framework for generating codelists which addresses the issues highlighted above. In particular, it provides a transparent and reproducible codelist which makes use of automation where possible to reduce the amount of input needed by clinical colleagues.

The proposed approach (shown in Fig. 1) for the Generalised Codelist Automation Framework (GCAF) illustrates how to develop codelists using automation to minimize the workload for clinicians. The different modules of the GCAF are described below, using DynAIRx to ground the description. For context, DynAIRx uses primary care data from CPRD to predict adverse drug reactions in those with MLTCs (full details in Section-4).

**Fig. 1:**
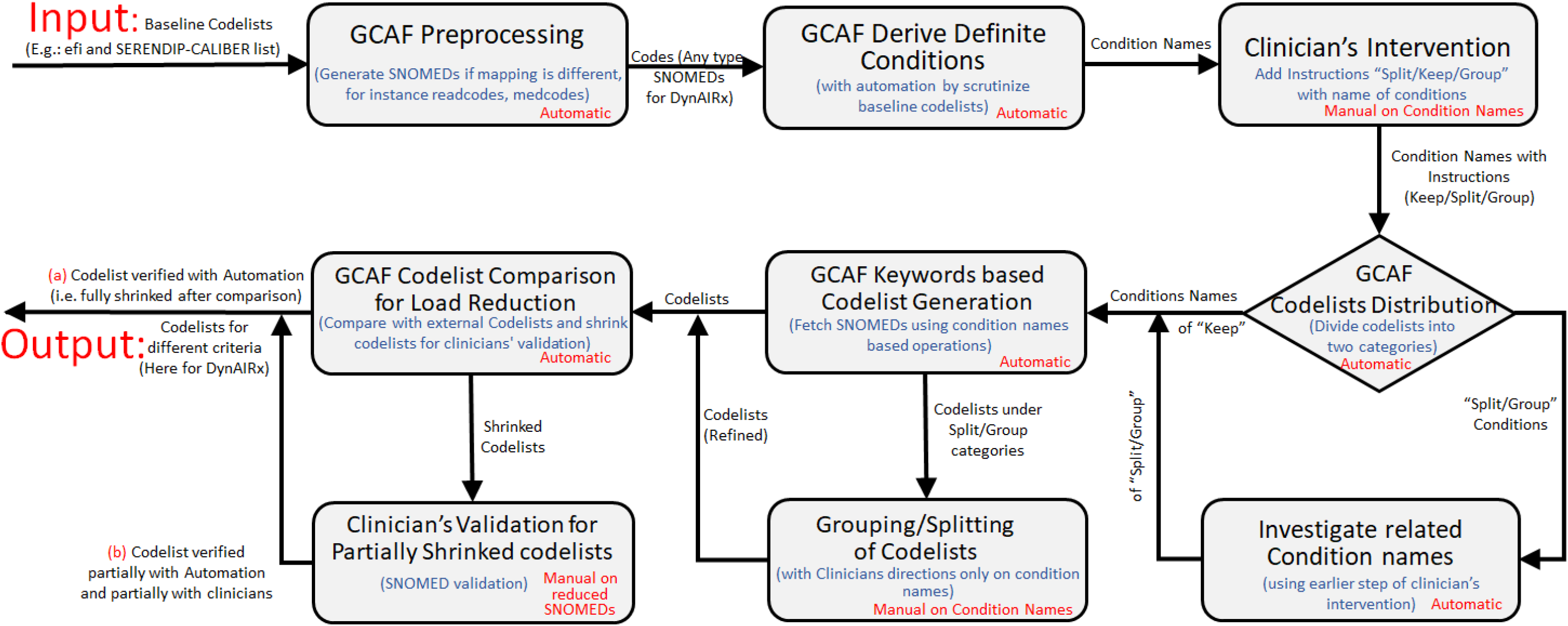
Generalised Codelist Automation Framework (GCAF).

To begin, we assume there will be existing codelists that are related to the use-case of the project as a starting point to build upon. In this case we begin using two codelists for MLTCs that have been clinical validated previously: eFI2 [45] and SERENDIP [46] (the latter based upon CALIBER).

#### GCAF Preprocessing

In the first step, mapping files from NHS TRUD, for example, are used to convert these initial lists containing Readcodes, Medcodes, SNOMEDs etc., into the required format. For DynAIRx, SERENDIP codes needed converting from Read v3 to SNOMED. Once mapped into a uniform ontology, these codelists are transferred to the next module.

#### GCAF Derive Definite Conditions

We commonly find that concepts can be given different names across codelists, including use of spaces, under-scores, capital letters, joining two names, and plurals etc., though we need to have consistency in the names for automation. Matching concepts across these preprocessed lists allows us to create a list of definitive conditions. The purpose of this module is to scan all input codelist, perform text operations on condition names, and generate a list of definitive conditions. For instance, name of conditions like Alcohol-related Brain Injury, Autoimmune liver Disease, Pulmonary hypertension, Chronic Obstructive Pulmonary Disease (COPD), Anaemia Folate Deficiency, Schizoaffective etc. The definitive list of conditions is then used in the next module “*Clinical Intervention*”.

#### Clinical Intervention

This intervention module is important in receiving guidance on which clinical concepts in the codelist need to be split and which to be grouped or merged based on the specific usecase of the project. For example, it may be necessary to split mental health into subsets for depression, anxiety, etc. This manual step is only working with the names of conditions rather than individual SNOMED codes at this stage. In DynAIRx, we capture *≈* 200 concepts, which makes it the largest MLTC codelist in the UK to the best of our knowledge. In this phase, clinician’s add comments like “keep”, “group”, and “split” for condition names. For instance, in the case of abdominal hernia, if clinicians’ indicate “keep”, then GCAF will leave all underlying SNOMED codes for this condition alone. Similarly in case of “Alcohol Problems Others” if clinicians’ suggest “Group” which indicates merging with similar name codelists, so GCAF will automatically find other condition names which consist of “*Alcohol Problems Others*” and then merge all SNOMED codes in one category with final condition name as “*Alcohol Problems Others*”. It could be possible a category need to be split into multiple categories like “Alcohol” can be classified into “*Alcohol related Brain Injury* “, “*Alcoholic Liver Disease*”, “*Alcohol Problems*” and other problems.

#### GCAF Codelists Distribution

On the basis of the clinicians’ comments, we distribute conditions into two types “keep” comments type and “Group/Split” type comments. This is just an automated decision phase, which helps in deciding which list of conditions can be processed directly by GCAF and which need more attention.

#### Investigate related condition names

In this module, we focus on those concepts where the clinical team indicated the need for merging or splitting to produce a potential list of concepts we need to capture. Finding condition names that need to be grouped and/or split is a fairly manual process but can largely be done without clinical oversight at this stage. For example, the SERENDIP codelist has concepts for “*Macular Degeneration*”, “*Visual Impairment and Blindness*” and “*Cataract* “ whilst eFI2 has just *“Visual impairment”*. The comment from clinicians was to split *“Visual impairment”* into the constituent parts. This modules takes care of finding similar texts using comments and generate list of related condition names. So for this specific example, our draft list of concepts for *Macular Degeneration* is “*Macular Degeneration*”, “*Cataract* “, “*Visual Impairment and Blindness*”, and finally “*Visual impairment* “ to catch non-specific SNOMED terms.

#### GCAF Keywords based Codelist Generation

This automated phase performs a keyword search across the preprocessed codelists (using terms from the previous step), fetching the associated SNOMED codes, and generating draft codelists for condition names agreed in the previous module. For simpler conditions this can often complete the majority of the codelist, whilst more complicated conditions are handled in the net module.

#### Grouping/Splitting of Codelists

In this module we focus on those concepts deemed more difficult, usually due to the need for splitting and grouping, using the draft list of concepts from the module “*Investigate related condition names*”. In this module clinicians agree upon the final divisions or grouping of categories by skimming through the draft codelists. In our example of *Macular Degeneration*, our intermediate categories were “*Macular Degeneration*”,”*Visual impairment* “, “*Cataract* “, and “*Visual Impairment and Blindness*”. After clinical feedback these conditions were split into *Cataract, Macular Degeneration, Blindness, Visual Impairment and Blindness, Visual Impairment Diabetic, Visual Impairment Macular, Visual Impairment Diabetic and Macular, Visual Impairment Diabetic and Cataract*, and *Visual Impairment Other*.

#### GCAF Codelist Comparison for Load Reduction

In this module, the codelist outputs from the last two modules are “shrunk” where possible using a trusted source where the concepts match our requirements. Using a trusted source to verify parts of the draft codelists can significantly reduce the amount of clinical effort needed in subsequent validation. In DynAIRx, we used the CALIBER [4] codelist for matching of codes and shrinking codelists where the clinical concepts matched. This process automatically verified > 90% of the codes, leading to a huge reduction in the amount of time needed by our clinical team. A full analysis is given in Section–5.

Specifically “**shrinking**” means automatically validating codes using a trusted source (CALIBER for DynAIRx), as such codelists have already been clinically validated. If the codelist gets “Fully Shrinked” i.e. 100% that means “all” codes were already present in the trusted sources and therefore don’t need validation from clinicians in the project. If it’s “Partially Shrinked” then some were validated via automation and a few codes need manual validation from clinicians. In the next section discussing DynAIRx, we find that this saves an enormous amount of clinical effort.

#### Clinician’s Validation of Partially Shrunken codelists

This final module of the GCAF requires clinical validation of the partially shrinked codelists. A secondary benefit of this phase is the final verification of new SNOMEDs by clinicians which can be trusted in future projects. The additional number of new codes added by DynAIRx, with comparison with existing codelists, is also detailed in Section–5.

It is important to note that “*GCAF Preprocessing* “, “*GCAF Derive Definite Conditions*”, “*GCAF Codelists Distribution*”, “*GCAF Keywords based Codelist Generation*”, “*GCAF Codelist Comparison for Load Reduction*”, and “*Investigate related Condition names*” modules are largely automated by GCAF and our software is available for use by researchers intending to develop codelists within reduced time for their usecase. We indicated same with Automatic/Manual labels in different modules of Fig.1. “*Clinical Intervention*” and “*Grouping/Splitting of Codelists* “ phases requires clinical guidance only for the name of conditions, there is no need to go through 1000s of SNOMED codes in these phases, and condition names could be few in number, depending upon the focus of the project. The “*Clinician’s Validation for Partially Shrunken codelists*” phase requires clinical feedback but the shrinking process dramatically reduces the amount of effort required. We show the workload reduction for DynAIRx in Section–5.

We implemented and managed this framework within our team using GitHub^4^. Different *GitHub branches* are used to integrate the different conditions like alcohol, pulmonary, heart diseases, etc. with one individual responsible for reviewing and merging branches. Various tasks are assigned and tracked by designating *GitHub Issues* for each team member. We are releasing our repository publicly with this paper, and recommend using GitHub (Issues, Branches, Comments, Documentation) for maintenance and development of the codelists of future projects.

## 4 Case Study: DynAIRx

The DynAIRx (Dynamic Artificial Intelligence for Medicines Optimisation) project aims to develop new, easy to use tools that support GPs and pharmacists to find patients living with multimorbidity (two or more long-term health conditions) who might be offered a better combination of medicines [47, 48]. DynAIRx uses structured EHR data from the Clinical Practice Research Datalink (CPRD^5^ [49–52]).

The National Health Service (NHS) introduced Structured Medication Reviews (SMRs), undertaken by GPs and pharmacists, with an aim to reduce the number of people taking potentially harmful drug combinations. However, there is no easy way of predicting who is most likely to benefit from a medication review. The DynAIRx project is developing tools to combine information from EHRs, clinical guidelines and risk-prediction models to ensure that clinicians and patients have the necessary information to prioritise and support SMRs.

DynAIRx focuses on multimorbidity [42] and polypharmacy[53, 54] within three key groups: (a) Older people with frailty, (b) People with co-existing mental and physical health problems, and (c) Other people with complex multimorbidity (≥ 4 long-term conditions). ‘Multimorbidity’ [55–60] is a priority for global health research’, and defined by the NIHR as the co-existence of two or more long-term conditions, each one of which is either (a) A physical non-communicable disease of long duration, such as a cardiovascular disease or cancer. (b) A mental health condition of long duration, such as a mood disorder or dementia. (c) An infectious disease of long duration, such as HIV or hepatitis C.

In the following subsections we detail the application of the GCAF for DynAIRx.

### 4.1 Baseline codelists

In this design we used two baseline codelists from previous projects: the eFI2 (expansion of the electronic frailty index (eFI)[45]), and the “SERENDIP codelist” from a project on relational pattern mining for multi-morbidity [46]. The eFI is a frailty indicator derived from routinely available primary care electronic health record data and designed to support the identification of elderly people living with frailty. “SERENDIP” is a modified subset of CALIBER. We also made use of the NHS Digital SNOMED CT Browser ^6^ for manually creating some of the conditions.

### 4.2 Simple Conditions for GCAF within DynAIRx

To begin, we start by preprocessing the baseline codelists (discussed in Section–4.1). In this step we convert all the codelists into SNOMED.

After mapping to SNOMED, we derive the specific condition names like *Epilepsy, Abdominal Hernia, Anxiety, Heart failure* etc. This list of conditions was passed to “Clinician’s Intervention” to add comments regarding to keep all SNOMEDs, or split into sub categories or group with any other categories. Clinician’s of DynAIRx indicated one of “keep”, “group”, or “split”. Those marked “keep” were deemed to be “Simple Conditions” which do not require further aggregation or dis-aggregation. GCAF generates automatic codelists for such conditions, and then automatically validate them through CALIBER codelists (see Fig. 1). After this automatic validation, there are a small number of SNOMED codes requiring clinical validation, drastically reducing the clincal effort required.

Using this approach, we generated 112 conditions which each consist of between 2 and 207 SNOMED codes (see Fig. 2 for details including the name of conditions and their corresponding number of SNOMED codes). We observed that we have 15 conditions which each consist of 80 to 207 number of codes, 40 codelists which consist of 30 to 80 codes, 20 codelists with ∼ 20 codes, 25 codelists with ∼ 10 codes, and 35 codelists consisting of 2 to 10 codes.

**Fig. 2:**
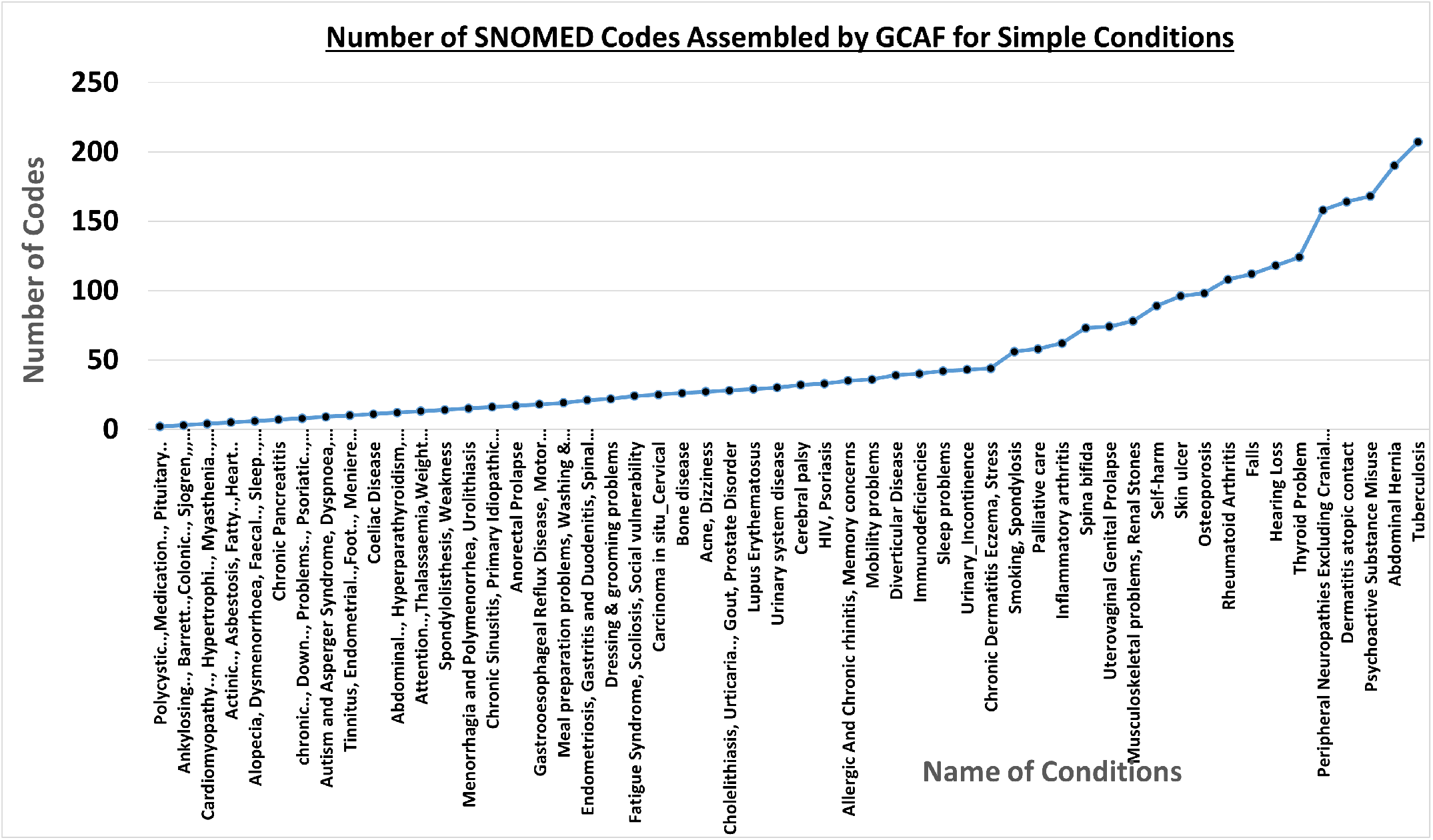
Simple conditions retrieved by GCAF along with the number of SNOMED codes they contain.

### 4.3 Complex Conditions for GCAF within DynAIRx

This subsection covers the conditions where clinicians recommended we group or split concepts, and with detailed instructions. Some examples of complex conditions are shown in Fig. 3. GCAF generates draft codelists for each of them using automation and then we manually merge or divide them following clinical recommendation. After this GCAF, shrinks and automatically validates codes using CALIBER (our trusted source) where possible and the remainder are validated by the clinical team. These codes get validated in short meetings, and the final codelists for DynAIRx are then agreed. Some examples of the journey when preparing difficult conditions are discussed below.

**Fig. 3:**
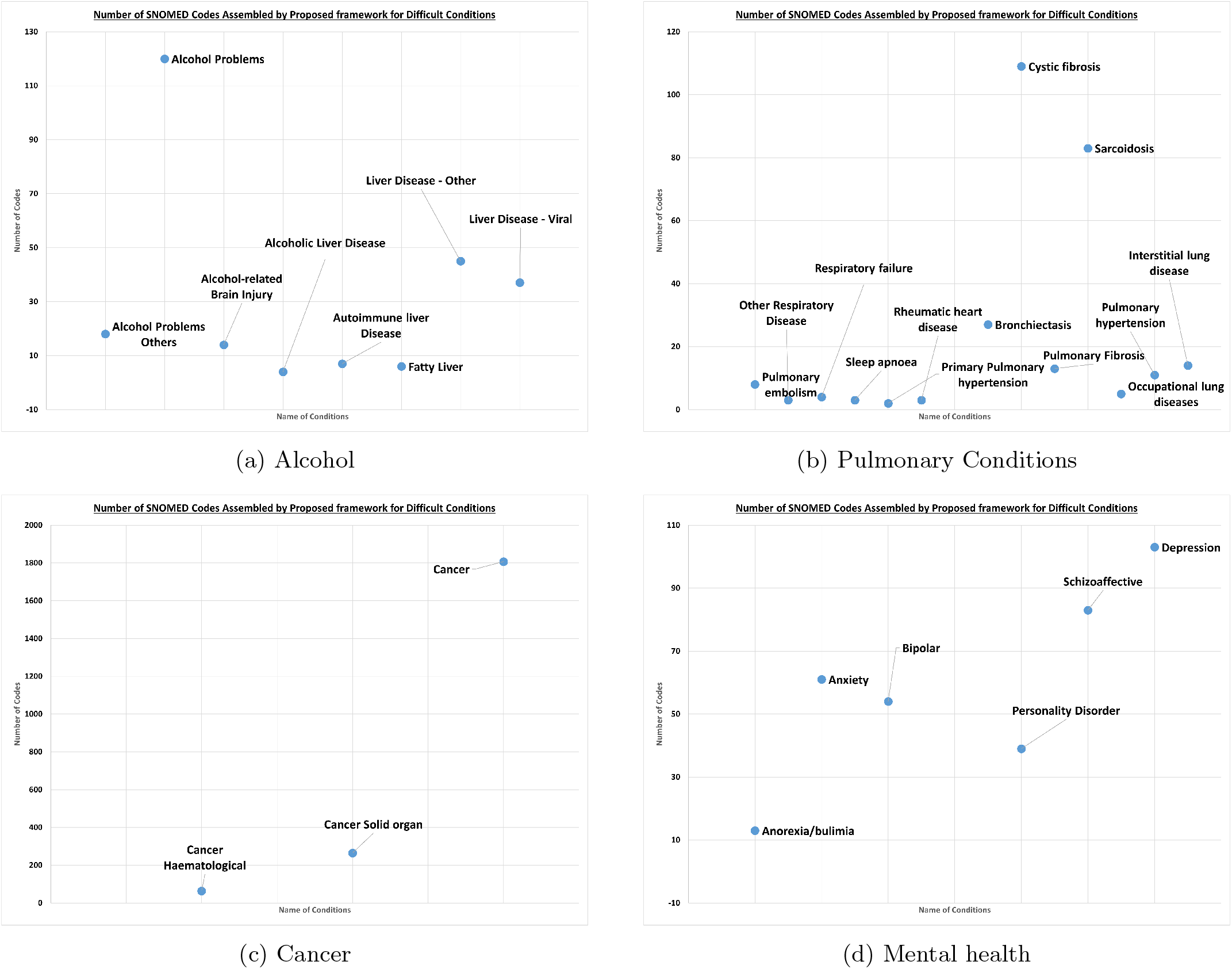
Complex condition analysis after following clinical recommendations.

#### Alcohol

Upon analysis of ‘Alcohol’ related conditions, we found the initial concepts: *Alcohol Problem, Alcoholic Liver Disease, Alcohol-related Brain Injury, Autoimmune liver Disease, Chronic Liver disease And Viral hepatitis, Oesophageal varices, Alcohol, Liver problems*, and *Fatty Liver*. We prepared draft codelists for these conditions using the GCAF modules and passed only the condition names to clinicians to review. Some example comments included: “*eFI list includes codes for alcoholic liver disease. separate and have 1) alcohol problems and 2) alcoholic liver disease and 3) alcohol brain injury. The current eFI list is missing some of the alcohol brain injury codes*”, and “*would prefer to break this down by cause (alcohol being one, viral being another)’* ‘. Using these guidelines we created final codelists of “Alcohol Problems”, “Alcohol-related Brain Injury”, “Alcohol Problems Others”, “Alcoholic Liver Disease”, “Autoimmune liver Disease”, “Fatty Liver”, “Liver Disease - Other”, “Liver Disease - Viral”. These were shrunk with the remainder undergoing clinical review Fig. 3a.

#### Cancer

For cancer, we initially generated codelists for “*Cancer Haematological* “ and “*Cancer Solid organ*” from SERENDIP and a generic “*Cancer* “ list from eFI2. After removing duplicates, this was shrunk and taken for clinical review. In this case we only removed a few rows of cancer using automation and the final number of codes for these conditions are shown in Fig. 3c.

#### Pulmonary Conditions

During analysis of pulmonary conditions, we initially drafted the following list of conditions: *Chronic Obstructive Pulmonary Disease (COPD), Respiratory disease, COPD, Asthma, Primary Pulmonary Hypertension, Pulmonary Fibrosis, Recurrent pulmonary embolus*, and *Secondary Pulmonary Hypertension*. Clinical feedback on this intial list led to the following comments: “in eFI, can check and compare the codelists? asthma and COPD and pulmonary fibrosis are distinct codelists, is this covering others?”, “compare codelists”, “need to compare codelists for consistency”, “need to compare codelists for consistency”, and “Keep this - not currently covered by eFI list”. Following these comments, the final list of conditions that were shrunk and sent for clinical review were “Chronic Obstructive Pulmonary Disease (COPD)”, “Pulmonary embolism”, “Other Respiratory Disease”, “Respiratory failure”, “Sleep apnoea”, “Primary Pulmonary hypertension”, “Rheumatic heart disease”, “Asthma”, “Bronchiectasis”, “Cystic fibrosis”, “Pulmonary Fibrosis”, “Sarcoidosis”, “Occupational lung diseases”, “Pulmonary hypertension”, and “Interstitial lung disease”. The size of these conditions is shown in Fig. 3b.

#### Mental Health

We invested a significant amount of time in properly capturing mental health conditions. Our initial draft list of conditions included *Learning Disability, Cognitive impairment, Dementia, Anxiety, Schizophrenia, depression*, and different *General mental health disorders*. We got lots of feedback including: “*Keep this - not currently covered by eFI list, might be some overlap with codes within cognitive impairment list - compare*”, “*include*”, “*compare to eFI cognitive impairment, dementia and memory problems list. happy to split into those 3 categories as per eFI* “ etc. After some clarification we were left with the following conditions “*Anorexia/bulimia*”, “*Anxiety* “, “*Bipolar* “,”*Dementia*”,”*Personality Disorder* “,”*Schizoaffective*”, and “*Depression*”. The number of codes for each mental health condition is shown in Fig. 3d.

Some of the other complex conditions requiring significant effort included tremor, fracture, headache, anaemia, stroke, bleed, angina, and ischaemic heart disease. Full details can be found by exploring our publicly available codelist repository (attached with proposed work), in particular the Github issues.

### 4.4 Shrinking of Codelists for Reduced Validation Requirement

All the complex and simple conditions went through the shrinking procedure prior to clinical review. In this phase we match the condition names with those of CALIBER to remove those SNOMED codes which were previously validated by CALIBER. For example: ***Bladder Dysfunction*** to the CALIBER lists ***kuan neuro bladder*** and **kuan pri bladder, *OCD*** to ***kuan ocd, Sickle cell anaemia*** to ***kuan sickle cell*** and ***kuan sickle trait, Gastritis and Duodenitis*** to ***kuan gastritis duodenitis*** etc. We then remove overlapping codes of all codelists, and sent the remaining codes (that are not present in CALIBER) for clinical review. An evaluation of the amount of workload this saved our clinical team is given in Section–5.1.

### 4.5 Clinical Validation Strategy used with DynAIRx

After we automatically verify most of the SNOMED codes using CALIBER, we put the remaining “shrunken codelists” to a clinical review. Within DynAIRx we followed a strict strategy for reviews.

#### Strategy for clinical review

Codelists which fit at least one of the criteria below need to be reviewed by at least two clinicians:

- Are a subset of a previously verified codelist but have not been clinically reviewed in any other context
- Are a combination of previously clinically verified codelists but have not been clinically reviewed in any other context
- Are for a condition without any existing published codelist (that has undergone clinical review)
- Involve test codes (e.g. diagnosis as a result of a biomarker; in the case that none of the above criteria are fulfilled only the test codes need to be reviewed)

Codelists will not need to be manually reviewed if none of these criteria are met, meaning that codelists which have been previously published and require no modification when following our clinical comments may be used in DynAIRx without further review. In this case, the source of the codelist will be clearly logged.

#### Review process

All decisions will be clearly logged in the corresponding code files used to generate the final codelists (primarily notebook files, stored in GitHub, codes provided with paper).

#### Ambiguous/boundary codes

Some codes might be mildly or highly suggestive of a condition but not exclusive to it. Some researchers may favour including this code (prioritising sensitivity of codelists) whilst others may choose to exclude it (prioritising specificity), hence the terminology of ‘boundary’ codes. In cases where clinical input suggests a code may be considered a boundary case, this will be clearly logged (through a column ‘Boundary case’ in the codelist CSV files which contains a value of 1 if a boundary code). Boundary codes must be reviewed by 2 clinicians, and each must give a preference for inclusion or exclusion (it is unlikely that this will be done in a blind or anonymous manner, since codelist review is likely to occur within a clincial panel). In the case where two clinicians disagree, a third will be used for a casting vote. Boundary codes which are chosen to be excluded will be noted in the corresponding code file used to generate the codelist.

#### Prevalence checking

Following completion of the codelists, the prevalence of conditions within the DynAIRx populations will be calculated and compared to published estimates of similar populations (where available). Should the estimates obtained using DynAIRx codelists be significantly different to those published estimates, codelists will be (re)reviewed. It is expected that DynAIRx prevalence estimates will be higher than those in the general population due to the multimorbid nature of the cohorts.

#### Patients and Public Involvement and Engagements (PPIE) Workshops

DynAIRx also conducted three workshops on Patients and Public Involvement and Engagement (PPIE). First in-person workshop (Feb 2024) was focused on aim of DynAIRx optimizing Structured Medical Reviews (SMR) using AI, involving six Work Packages (WP), focused on SMRs, AI Prediction, Causal Inference, Natural Language Processing (NLP), Visualization, and focusing on PPIE feedback. The second meeting (online in April 2024) was about listening of two PPIE members, clarifying their doubts, and noting things they want us to incorporate in our communication groups. The third workshop (in-person, June 2024) was held in the Civic Health Innovation Labs (CHIL), with the University of Liverpool about presenting AI work of clustering of patients trajectories for optimizing medications and generation of codelists along with proposed framework. As outcomes of these workshops mainly cover the priority of conditions, side effects, the burden of long-term prescriptions on patients, and feedback from public advisors using AI for medication optimizations, more details of them are out of the scope of this paper.

## 5 Findings and Discussion

In this section we demonstrate the benefits of using this framework within DynAIRx across four experiments.

1. We demonstrate the efficiency of GCAF by quantifying the reduction in number of SNOMED codes requiring clinical review.
2. We compare existing codelists applicable to MLTC research with the DynAIRx codelist, discussing their strengths and weaknesses, and the number of conditions and codes.
3. We investigate how many “*new* “ codes DynAIRx codelist adds over CALIBER codelist in each *condition*, which makes DynAIRX codelist more comprehensive for public use. All of these new codes have been through our clinical review process.
4. Finally, we discuss the time investment of clinicians, and the phases of review. Appendix-A provides the list of condition names that are covered in DynAIRX and the codelists generated by GCAF.

### 5.1 Results on Shrinking of lists

An analysis of the percentage of codes that could be shrunk during the codelist development is presented in Fig. 4. Here, for each condition, shrinking percentages show the proportion of codes that could be automatically verified using CALIBER. To simplify the figure, only 90 conditions are shown in Fig.4a, 4b, 4c, and 4d. Specifically, Fig.4a consisted of codelists with less than 10 codes, and we see that automation saved 100% of the work for most of the codelist except for Alcoholic Liver Disease 66.67%, Fatty Liver 80%, and Sick Sinus Syndrome and conditions related to Cardiomyopathy at 0%. In Fig.4b most of the conditions were 100% shrunk using GCAF, thus saving clinical time, apart from Chronic Tinnitus 0% Abdominal Aortic Aneurysm 36.35% and Pulmonary Fibrosis 91.67%. Similarly Fig.4c and 4d contain many conditions which were fully shrunk and therefore did not need further review. Overall, we observe that many conditions like Fragility fracture, Dermatitis atopic contact, Self-harm, Musculoskeletal problems, HIV, Anterior and Intermediate Uveitis were automatically constructed by GCAF (Fig. 1). Some of the more complicated conditions including Alcohol Problems were 54.62% automatic, similarly Psychoactive Substance Misuse at 84.43%, Cataract at 87.38%, Fatty Liver at 80.00%, and Abdominal Aortic Aneurysm ay 36.36% etc. A number of conditions such as Polycythaemia vera, Sick sinus Syndrome, and Chronic Tinnitus were newly introduced within DynAIRx, and so require full clinical review. Overall GCAF automation validated > 80% of the SNOMED codes, leaving < 20% requring clinical time for review.

**Fig. 4:**
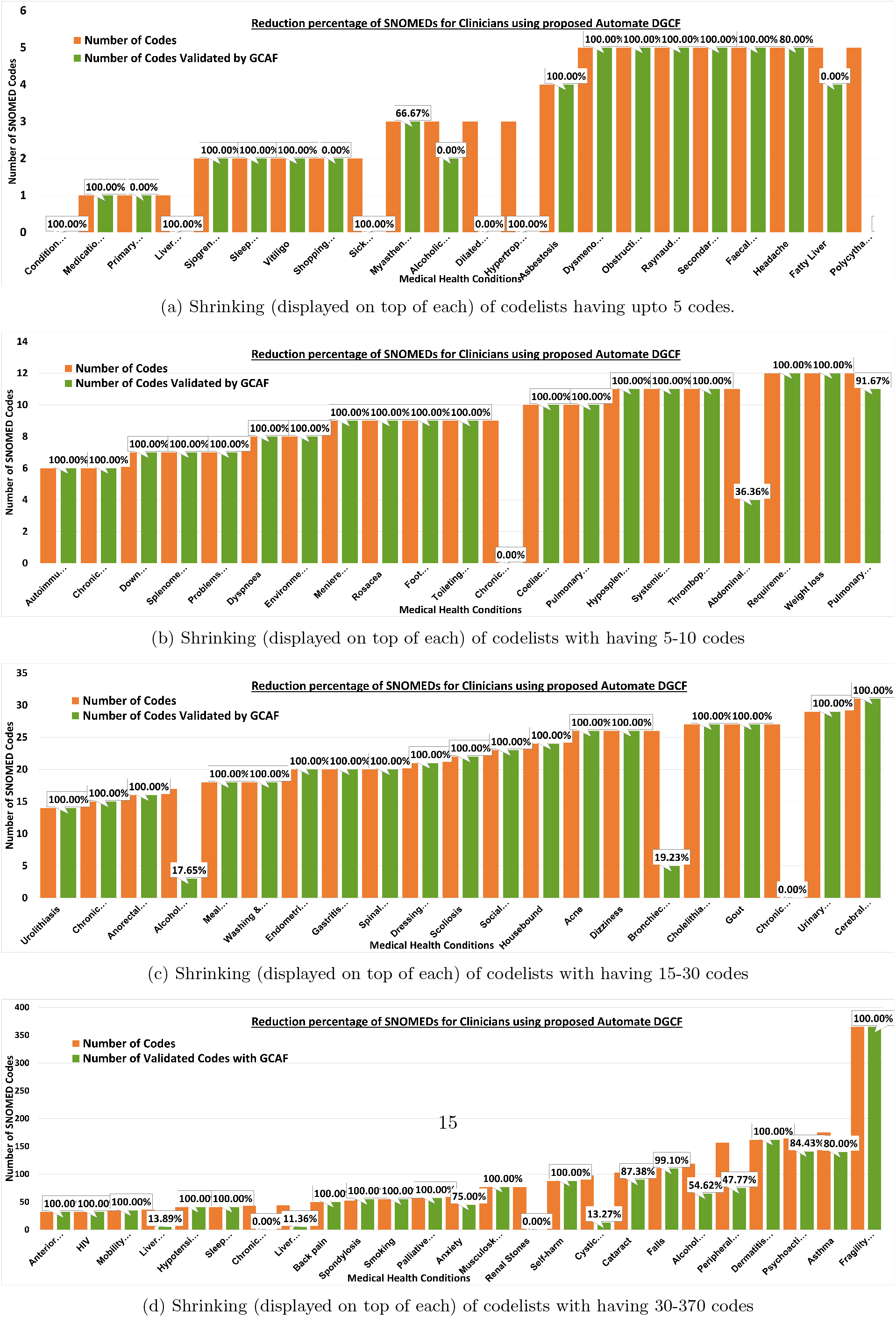
Reduction percentage (displayed on top) of SNOMED codes for clinicians with and without using automation in GCAF. We separated the conditions across 4 figures to display the reduction percentage clearly.

### 5.2 Comparison with Existing Codelists

Table 1 presents a comparison of existing codelists with DynAIRx. We compare the DynAIRx codelists to CALIBER, eFI2, AI-Multiply, and Optimal. Details of the origin of these codelists is presented in Section–2.3. All these codelists are from recent projects in the area of multimorbidity. We compared codelists based on the number of conditions (d) Shrinking (displayed on top of each) of codelists with having 30-370 codes covered, number of codes, number of conditions related to MLTC, and number of total codes related to MLTC. Note that CALIBER contains a number of codelists which are specific to COVID-19. In DynAIRx codelists, we did grouping (merging) of different conditions and removed conditions that are not directly related to MLTC so codelist count is less but number of conditions are more than existing codelists. Please note, codelists that we are not considering in DynAIRx can be found in separate folder within codelists (attached in supplementary material of this paper).

**Table 1:**
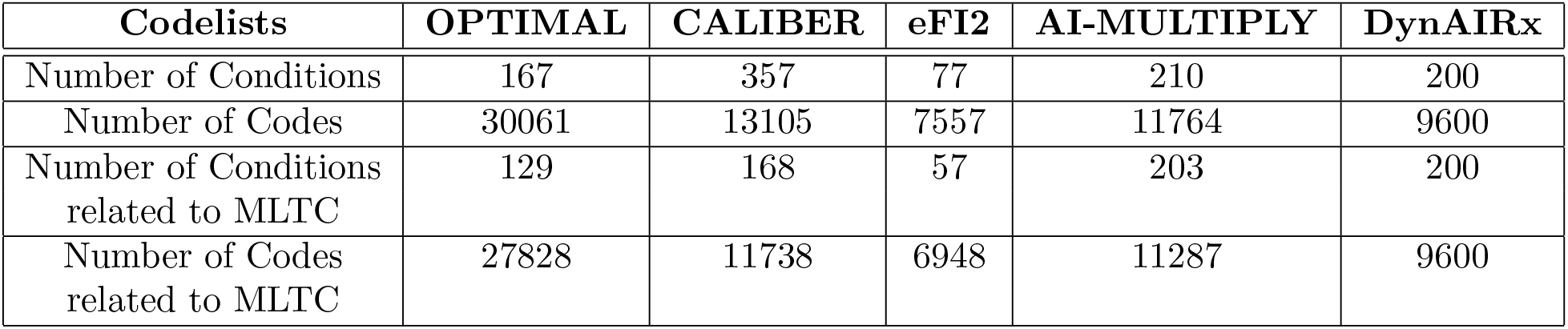
Comparison of existing codelists used within MLTC research and codes (all based on SNOMED).

We find that Optimal, and AI-MULTIPLY are large codelists but consist of many conditions which are not technically LTCs. Table 1 summarises the differences between the codelists.

### 5.3 DynAIRx Added New Codes

As previously discussed, the DynAIRx codelist aims to be comprehensive within the UK for research involving MLTCs. In Table 2 we show the number of new SNOMED codes added in various conditions compared to CALIBER. We find that DynAIRx codelist adds a number of new SNOMED codes for conditions including Renal Stones, Peripheral Neuropathy, Alcohol Problems, Chronic Dermatitis Eczema, and Cystic fibrosis etc. The full list of *≈* 200 conditions covered by DynAIRx and released in our repository can be seen in Appendix-A.

**Table 2:** Conditions with new SNOMED codes added in DynAIRx compared to CALIBER codelists.

| Condition Name | Number of New Codes Added | Condition Name | Number of New Codes Added |
| --- | --- | --- | --- |
| Abdominal Aortic Aneurysm | 8 | End Stage Renal Disease | 37 |
| Alcohol Problems | 55 | Fatty Liver | 2 |
| Alcohol Problems Others | 15 | Falls | 2 |
| Alcoholic Liver Disease | 2 | Hypertension | 56 |
| Anaemia B12 Deficiency | 13 | Hypertrophic Cardiomyopathy | 4 |
| Anaemia Folate Deficiency | 5 | Hypotension/Syncope | 23 |
| Anaemia Haemolytic | 13 | Liver Disease - Other | 40 |
| Anaemia Iron Deficiency | 8 | Liver Disease - Unknown | 2 |
| Anaemia Other | 25 | Liver Disease - Viral | 32 |
| Anxiety | 16 | Migraine | 7 |
| Asthma | 36 | OCD | 6 |
| Back pain | 78 | Osteoporosis | 39 |
| Benign | 33 | Peptic ulcer Disease | 34 |
| Bronchiectasis | 22 | Peripheral Neuropathies | 83 |
| Cataract | 14 | Polycystic Ovarian Syndrome | 2 |
| Chronic Dermatitis Eczema | 44 | Polycythaemia vera | 6 |
| Chronic Tinnitus | 10 | Psychoactive Substance Misuse | 27 |
| Chronic Urticaria | 28 | Pulmonary Fibrosis | 2 |
| CKD | 15 | Renal Stones | 78 |
| Cystic fibrosis | 86 | Sick sinus Syndrome | 3 |
| Depression | 62 | Sickle cell anaemia | 17 |
| Dilated Cardiomyopathy | 4 | Thyroid Problem | 79 |

### 5.4 Clinical Validation

As discussed in Section–2, the limitations of existing codelist development methods include time investment and limited use of automation leading to inefficiency and potential for error. Often this requires a huge investment of time from clinical colleagues, who also have clinical commitments, making codelist development a lengthy process. To demonstrate the huge potential for saving time when using GCAF, we list all the clinical engagement required for the DynAIRx codelist in Table 3.

**Table 3:**
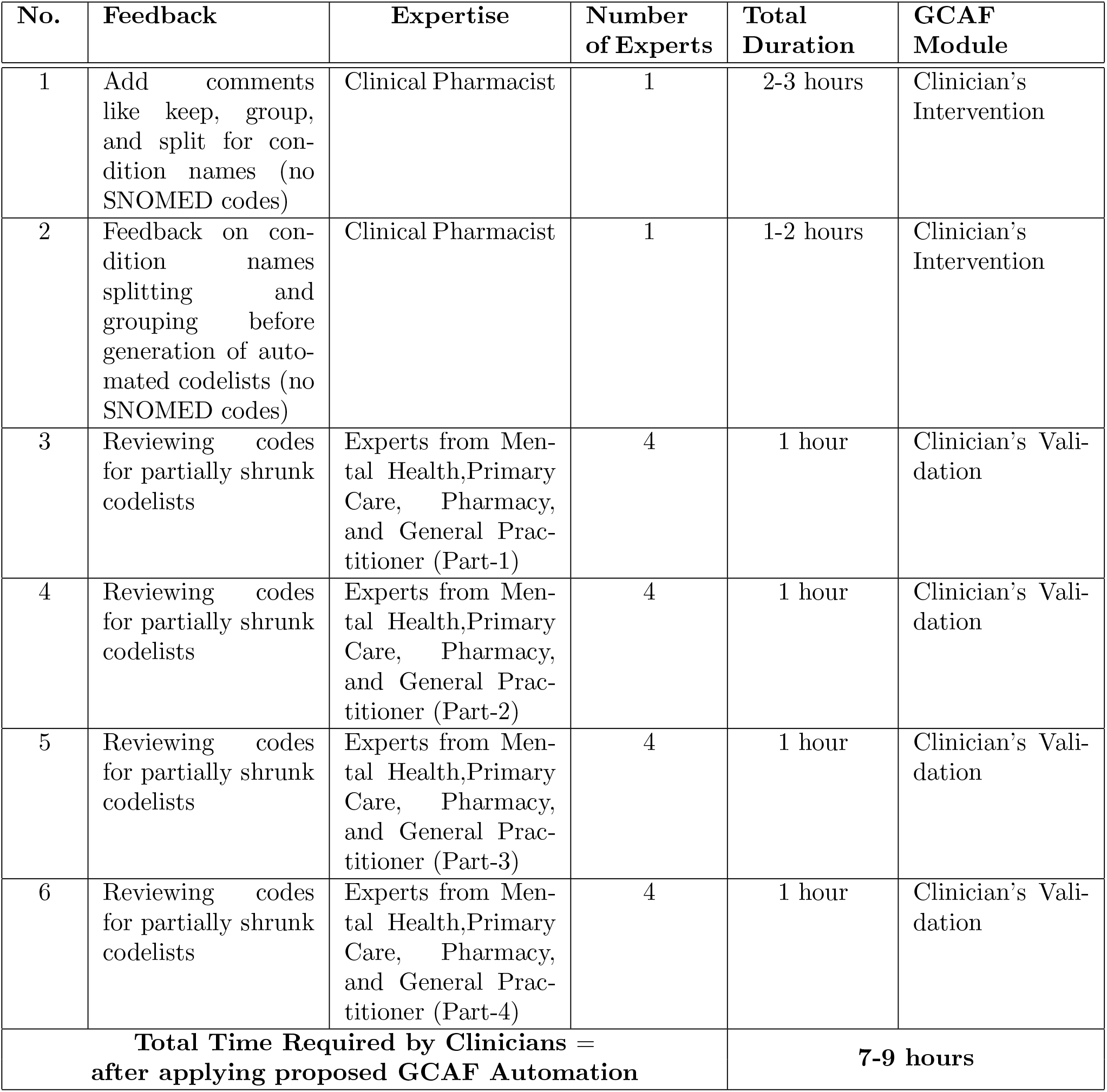
Type and time investment of clinicians for the development and validation of codelists generated in DynAIRx, using GCAF.

| No. | Feedback | Expertise | Number of Experts | Total Duration | GCAF Module |
| --- | --- | --- | --- | --- | --- |
| 1 | Add comments like keep, group, and split for condition names (no SNOMED codes) | Clinical Pharmacist | 1 | 2-3 hours | Clinician's Intervention |
| 2 | Feedback on condition names splitting and grouping before generation of automated codelists (no SNOMED codes) | Clinical Pharmacist | 1 | 1-2 hours | Clinician's Intervention |
| 3 | Reviewing codes for partially shrunk codelists | Experts from Mental Health, Primary Care, Pharmacy, and General Practitioner (Part-1) | 4 | 1 hour | Clinician's Validation |
| 4 | Reviewing codes for partially shrunk codelists | Experts from Mental Health, Primary Care, Pharmacy, and General Practitioner (Part-2) | 4 | 1 hour | Clinician's Validation |
| 5 | Reviewing codes for partially shrunk codelists | Experts from Mental Health, Primary Care, Pharmacy, and General Practitioner (Part-3) | 4 | 1 hour | Clinician's Validation |
| 6 | Reviewing codes for partially shrunk codelists | Experts from Mental Health, Primary Care, Pharmacy, and General Practitioner (Part-4) | 4 | 1 hour | Clinician's Validation |
| **Total Time Required by Clinicians = after applying proposed GCAF Automation** |  |  |  | **7-9 hours** |  |

We see a huge reduction in the amount of clinical time required, particularly given the large size and scope of the DynAIRx codelist. The validation of the shrunk lists required only four 1-hour meetings with our team of clinicians. Prior to these meetings, the DynAIRX codelists also involved two meetings and a dozen emails to clarify the comments from the “Clinical Intervention” GCAF module.

Using the partially shrunk codelists that come from the automated part of GCAF, the four meetings for clinical validation were extremely efficient. In these four clinical meetings we covered 45 codelists in Phase-1, 24 in Phase-2, 17 in Phase-3, and 25 in Phase-4. The difference in coverage over time is because we primarily focused on the “easy” conditions in the first meeting and progressively moved to the more complicated ones (see Section–4.2 and Section–4.3). As previously shown, the automated shrinking process managed to reduce the number of codes needing manual review substantially allowing for hugely efficient meetings. After each meetings, we performed the minimal changes that were required and finalised each condition. The completed codelists are accessible in the publicly available repository for this work. Details of meetings and validation strategy can be found in Section–4.5 and Table 3. Our clinicians/experts (details can be found on our website ^7^) cover a variety of clinical specialities including pharmacology, mental health, geriatrics, psychiatry, internal medicine, and general practitioner.

## 6 Conclusion and Future Directions

In this work, we explored existing codelist development methodologies and barriers to having transparent, reproducible codelists. We found limited research that lived up to these ideals, with many not sharing their codelists, and others having opaque processes for codelist development. These are critical issues, as codelists need to be adapted for different research questions and cannot always be used as-is. The GCAF framework we propose addresses these issues and results in publicly accessible SNOMED codelists and software (Python toolkit) to aid codelist development. The primary advantage of this approach is the increased use of automation and using trusted sources to reduce the workload, which leads to to reduction in human error and considerable time saved (particularly the time investment required by clinical experts).

In addition to this, we provide a case study applying this approach to the DynAIRx project. In this case study we provide details of how to apply the framework in practice. We also provide evaluation of the development process which show the benefit of automation, strategies for having efficient clinical meetings, and a comparison against other contemporary codelists in the multimorbidity area. We conclude that by applying the proposed framework, a codelist of approximately *≈* 9500 codes requires only 7-9 hours of clinical time, with over > 80% of the codelists validated beforehand using automation from trusted sources. Finally, we release the codelists from DynAIRx (covering *≈* 200 conditions) and the software used to speed-up the development process for future researchers.

There are a variety of related topics in codelist development which deserve attention in future research. These include harmonization and interoperability between the various healthcare databases used across the UK (which currently require a variety of clinical ontologies), methods for maintaining codelists over time to avoid temporal drift, tools which help researchers to search, combine, and modify codelists from previously published research [61] (which may now be feasible due to advances in cutting-edge large language models), and standardization of codelists to enable reuse where possible. The development and release of automated frameworks such as GCAF are the first step in fully realising the capability of routinely collected healthcare in research.

## Data Availability

All data produced in the present study are available upon reasonable request to the authors

## Acknowledgements

DynAIRx has been funded by the National Institute for Health and Care Research (NIHR) Artificial Intelligence for Multiple Long-Term Conditions (AIM) call (NIHR 203986). MG is partly funded by the NIHR Applied Research Collaboration North West Coast (ARC NWC). This research is supported by the NIHR ARC NWC. The views expressed in this publication are those of the author(s) and not necessarily those of the NIHR or the Department of Health and Social Care. IB is funded by an NIHR Senior Investigator award (NIHR205131).

## Author Contributions

Conception & Design: SR, AA, LW; Data acquisition: AA (180+ conditions), HC (20 conditions), MA (20 conditions), MO (10 conditions), SR (Guidance on all); Proposed Framework and Analysis: AA, LW, SR; Clinical Validation: LW, AAW, ES, AC, JYT, FSM, SR; Writing: AA, IB, SR, MS, RAR; Final approval: all.

## Appendix A Appendix A: DynAIRx Codelist Stats

Please find the list conditions covered by the DynAIRx codelist in Table A1 using the Generalised Codelist Automation Framework (GCAF).

**Table A1:** All conditions covered by the DynAIRx codelist.

| DynAIRx Codelist Condition Names |  |  |  |
| --- | --- | --- | --- |
| Acne | Dizziness | Myasthenia Gravis | Social vulnerability |
| Activity limitation | Down Syndrome | Obstructive Sleep Apnoea | Spina bifida |
| Alcoholic Liver Disease | Dressing & grooming problems | Occupational lung diseases | Spinal stenosis |
| Alcohol-related Brain Injury | Dysmenorrhoea | Oesophageal varices | Splenomegaly |
| Anaemia B12 Deficiency | Dyspnoea | Osteoarthritis | Spondylolisthesis |
| Anaemia Folate Deficiency | End Stage Renal Disease | Osteoporosis | Spondylosis |
| Anaemia Haemolytic | Endometriosis | Palliative care | Systemic Sclerosis |
| Anaemia Iron Deficiency | Environment problems | Parkinson and Tremor | Thalassaemia |
| Anaemia Other | Erectile Dysfunction | Peptic ulcer Disease | Thrombophilia |
| Anorectal Prolapse | Faecal incontinence | Peripheral Neuropathies | Thyroid Problem |
| AnorexiaBulimia | Falls | Personality Disorder | Toileting problems |
| Anterior and Intermediate Uveitis | Fatty Liver | Pituitary adenoma | Urinary system disease |
| Anxiety | Female genital Prolapse | Polycystic Ovarian Syndrome | Urinary Incontinence |
| Asbestosis | Fibroids | Polycythaemia vera | Urolithiasis |
| Asthma | Foot problems | Polymyalgia Rheumatica | Vitiligo |
| Autoimmune liver Disease | Fracture | Primary Idiopathic Thrombocytopaenia | Washing & bathing problems |
| Autonomic dysfunction | Fragility fracture | Primary Pulmonary hypertension | Weakness |
| Back pain | Gastritis and Duodenitis | Problems managing finances | Weight loss |
| Benign | Glaucoma | Prostate Disorder | Abdominal Aortic Aneurysm |
| Bipolar | Gout | Psoriatic Arthropathy | Abdominal Hernia |
| Bladder Dysfunction | Headache | Psychoactive Substance Misuse | Actinic keratosis |
| Bone disease | Hearing Loss | Pulmonary Fibrosis | Allergic And Chronic rhinitis |
| Bronchiectasis | Heart Failure | Pulmonary hypertension | Ankylosing Spondylitis |
| Cancer | Heart block | Raynaud Syndrome | Attention Deficit Hyperactivity Disorder |
| Cancer Haematological | HIV | Renal and Bladder Stones | Autism and Asperger Syndrome |
| Cancer Solid organ | Housebound | Renal Stones | Barrett Oesophagus |
| Cataract | Hyperparathyroidism | Requirement for care | Carcinoma in situ Cervical |
| Cerebral palsy | Hypertension | Respiratory failure | Chronic Tinnitus |
| Cholelithiasis | Hypertrophic Cardiomyopathy | Rheumatic heart disease | Colonic polyp |
| Chronic Dermatitis Eczema | Hyposplenism | Rheumatoid Arthritis | Endometrial Hyperplasia and Hypertrophy |
| Chronic Obstructive Pulmonary Disease (COPD) | Hypotension syncope | Rosacea | Gastrooesophageal Reflux Disease |
| Chronic Pancreatitis | Immunodeficiencies | Sarcoidosis | Inflammatory arthritis |
| Chronic Sinusitis | Interstitial lung disease | Schizoaffective | Learning Disability |
| Chronic Urticaria | Liver Disease - Other | Scoliosis | Menorrhagia and Polymenorrhea |
| Chronic constipation | Liver Disease - Viral | Secondary Polycythaemia | Multiple Sclerosis |
| CKD | Lupus Erythematosus | Seizure Disorders | OCD |
| Coeliac Disease | Macular Disorders | Self-harm | Psoriasis |
| Complex pain syndrome | Meal preparation problems | Shopping problems | Skin ulcer |
| Cystic fibrosis | Medication management problems | Sick sinus Syndrome | Stress |
| Dementia | Meniere Disease | Sickle cell anaemia | Subdural Haematoma no-traumatic |
| Depression | Migraine | Sjogren Disease | Uterovaginal Genital Prolapse |
| Dermatitis atopic contact | Mobility problems | Sleep apnoea |  |
| Dilated Cardiomyopathy | Motor Neurone Disease | Sleep problems |  |
| Diverticular Disease | Musculoskeletal problems | Smoking |  |

## Footnotes

1 https://www.england.nhs.uk/long-read/population-health-management/

2 https://www.opencodelists.org/

3 www.liverpool.ac.uk/dynairx

4 https://github.com/

5 https://www.cprd.com/

6 https://termbrowser.nhs.uk/?

7 https://www.liverpool.ac.uk/dynairx/our-people/

